# Neurorehabilitation of the upper extremity – Immersive virtual reality vs. robot-assisted training. A comparative study

**DOI:** 10.1101/2023.09.19.23295411

**Authors:** Kira Lülsdorff, Frederick Benjamin Junker, Bettina Studer, Heike Wittenberg, Heidrun Pickenbrock, Tobias Schmidt-Wilcke

## Abstract

**Background:** Severe paresis of the contralesional upper extremity is one of the most common and debilitating post-stroke impairments. The need for cost-effective high-intensity training is driving the development of new technologies, which can complement and extent conventional therapies. Apart from established methods using robotic devices, immersive virtual reality (iVR) systems hold promise to provide cost-efficient high-intensity arm training.

**Objective:** We investigated whether iVR-based arm training yields at least equivalent effects on upper extremity function as compared to a robot-assisted training in stroke patients with severe arm paresis.

**Methods:** 52 stroke patients with severe arm paresis received a total of ten daily group therapy sessions over a period of three weeks, which consisted of 20 minutes of conventional therapy and 20 minutes of either robot-assisted (ARMEOSpring®) or iVR-based (CUREO®) arm training. Changes in upper extremity function was assessed using the Action Research Arm Test (ARAT) and user acceptance was measured with the User Experience Questionnaire (UEQ).

**Results:** iVR-based training was not inferior to robot-assisted training. We found that 84% of patients treated with iVR and 50% of patients treated with robot-assisted arm training showed a clinically relevant improvement of upper extremity function. This difference could neither be attributed to differences between the groups regarding age, gender, duration after stroke, affected body side or ARAT scores at baseline, nor to differences in the total amount of therapy provided.

**Conclusion:** The present study results show that iVR-based arm training seems to be a promising addition to conventional therapy. Potential mechanisms by which iVR unfolds its effects are discussed.

**Registry number:** DRKS00032489

## Introduction

Paresis of the contralesional upper extremity is one of the most common and debilitating impairments in the post-stroke condition. An estimated 77% of all first-time stroke survivors suffer from arm paresis ^1^. The probability of recovery has been shown to depend on the severity of the paresis at onset: While up to 70% of patients with a mild to moderate arm paresis regain some dexterity within the first six months post-stroke ^2^, up to 62% of patients with a severe arm paresis do not recover at all ^3^. The presence of a severe arm paresis has a strong negative impact on patients’ activities of daily living ^4,5^ and quality of life^6,7^.

Upper limb neurorehabilitation comprises not only mobilization and strengthening exercises aiming at the improvement of single and multiple joint movements, but also specific training of goal directed movements such as reaching and grasping. There is strong evidence that physical therapy post-stroke is most effective when applied with high intensity and repetition ^7–9^, and not surprisingly, increased amounts of therapy time are associated with better functional recovery and participation ^7,10–12^.

The need for cost-effective, high-intensity neurorehabilitation, together with an increasing lack of physio-and/or occupational therapists, is driving the development of technologies which have the potential to complement conventional therapy without increasing the need for personnel. One way to enhance the intensity of arm training and increase the number of movement repetitions is the implementation of robotic devices, which provide assistance of the paretic arm as well as direct visual feedback on movement performance ^7,13^. The anti-gravity support provided by robotic devices allows a wider range of movements and a higher number of repetitions, resulting in higher movement quality ^9,14^. Robot-assisted arm trainings have been shown to improve motor control of the paretic shoulder and elbow in both subacute and chronic stroke patients ^15^ and to be particularly beneficial for patients with severe motor impairments in the subacute phase after stroke ^16,17^. Especially exoskeletons such as the ARMEOSpring®, which provide anti-gravity support to the arm while guaranteeing high movement control of the arm and wrist, have been found to reduce upper limb impairment ^18^.

However, major disadvantages of robotic devices are the costs for acquisition and maintenance as well as the lack of mobility ^19^. Immersive virtual reality (iVR)-based trainings could potentially provide a cost-efficient, mobile alternative. Virtual reality (VR) is defined as a computer-simulated environment, with which the user can interact. In contrast to non-immersive VR, which is usually displayed on two dimensional computer screens, as for example in case of the ARMEOSpring®, iVR is most commonly displayed on HMDs (head mounted displays), which allow stereoscopic vision and give the user the illusion to be surrounded by a three-dimensional, computer-animated environment, with which he or she can interact in a seemingly physical way ^20^.

Systematic reviews and meta-analyses suggest that non-, semi- and immersive VR-based arm trainings have beneficial effects on motor function, range of motion and activities of daily living in stroke patients with arm paresis when provided in addition to conventional therapy ^21–23^. However, to date there are only a handful of randomized-controlled studies addressing the efficacy of iVR-based trainings in comparison to conventional therapy ^24–28^ in the treatment of severe arm paresis. Moreover, to our knowledge, there are no randomized-controlled studies on a direct comparison of iVR-based and robot-assisted arm trainings, which would allow a statement on whether iVR-based trainings truly offer an equivalent, yet mobile, alternative to robot-assisted trainings.

In the present study, we compared the efficacy of a newly developed iVR-based unilateral arm training for upper limb motor recovery after stroke (CUREO®) with the well-established robot-assisted ArmeoSpring® arm training. We hypothesized that iVR-based arm training would be non-inferior to the robot-assisted therapy. Both training methods were provided as adjuncts to standard rehabilitation in a group of subacute stroke patients with severe hemiparesis. As secondary outcome parameter, we investigated user acceptance.

## Methods

### Participants and study procedure

Sixty patients between 40 and 90 years were recruited on-site at the St. Mauritius Therapy Clinic (Düsseldorf – Germany). All patients had experienced a stroke between 13 days and 1 year before the study was conducted, resulting in a paresis of either the left or right arm. Exclusion criteria included pre-existing disabilities in hand function of the affected side, neurodegenerative or inflammatory diseases of the central nervous system, and non-correctable visual impairments. Patients with a complete hemiplegia of the arm were excluded; only patients who were able to lift their affected arm onto a table were eligible for study participation.

The study was conducted in accordance with the Declaration of Helsinki and had been approved by the Ethics Committee of the Ärztekammer Nordrhein (serial number: 2020079). Participants gave written informed consent prior to study enrolment.

The initial screening included a cognitive screening of the St. Mauritius Therapy Clinic, in which a chain of actions had to be performed by the patients (sharpen a pencil, write their name and the current date on a piece of paper, punch, fold and envelope paper, write their address on the envelope). Patients were only included when they were able to follow the instructions. Moreover, the Box and Block Test ^29^ for unilateral gross manual dexterity was performed. Patients were included when they were unable to lift more than ten blocks from one compartment to the other within 60 seconds. This was done to only include patients with a severe paresis of the upper extremity.

After initial screening, patients were randomly assigned to either the robot-assisted (n = 31) or iVR-based arm training (n = 29). Randomization was performed using the Minirand-package in R for minimized block randomization ^30^ and took place prior to the initial Action Research Arm Test (ARAT) (see Figure 1: ARAT #1). Three patients in the robot-assisted arm training and five patients in the iVR-based arm training did not finish this first assessment and were excluded, leaving 28 patients in the robot-assisted and 24 patients in the iVR group. Subsequently, the training period (intervention, for details see below) was initialized. Two patients of the robot-assisted group and five patients in the iVR group aborted the intervention (drop-outs). Subsequent ARAT scores were assessed from 26 patients after robot-assisted and 19 patients after iVR-based arm training (see Table 1 for demographic data and ARAT scores and Figure 1 for study procedure). Finally, reports on users’ experiences could be obtained from 23 patients after robot-assisted and 17 patients after iVR-based arm training using the User Experience Questionnaire (UEQ) ^31^.

**Figure 1:**
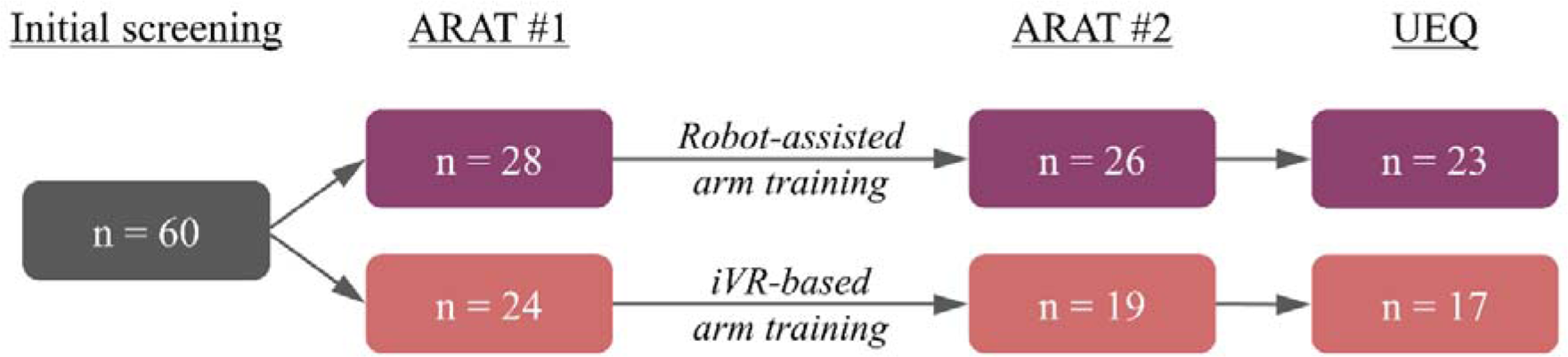
Study procedure with the corresponding number of patients for robot-assisted (violet) and iVR-based arm training (red).

**Table 1:**
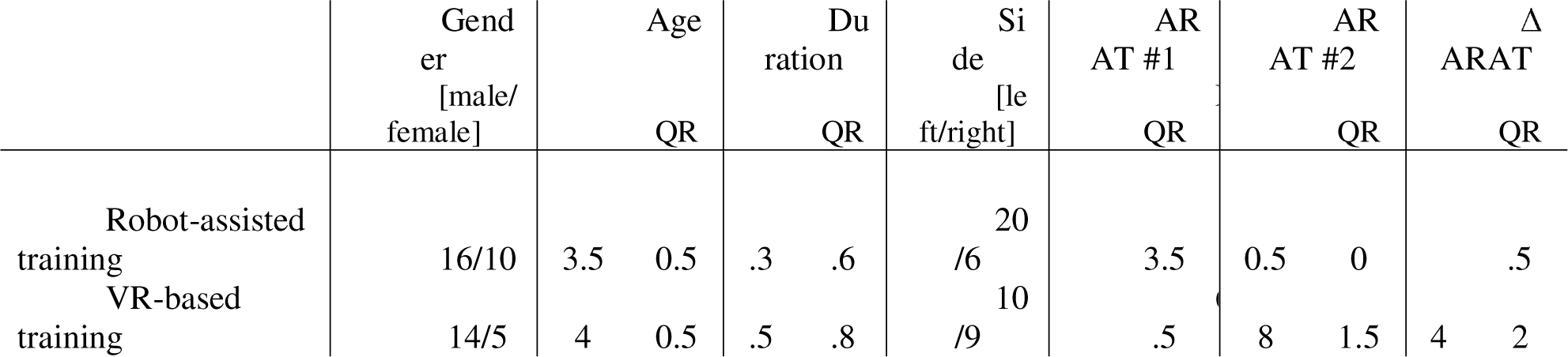
Demographic data and ARAT scores. Description of demographic data and ARAT scores before and after both interventions as median scores (M) with corresponding interquartile ranges (IQR). Age and duration after stroke are presented in years.

### Outcome measures

#### Action Research Arm Test (ARAT) – primary outcome variable

The Action Research Arm Test (ARAT) ^32^ is an observational rating scale of upper extremity function, which consists of 19 items grouped into four subscales: grasp, grip, pinch and gross movement. All items have a time limit of 60 seconds; however, if the patient is able to perform the test but requires more time, completion of the test is acknowledged. The items are scored on a four-level ordinal scale ranging from zero (unable to perform the test), one (partial performance of test), two (completes test in more than 60 seconds) to three (normal test performance). Total scores range from 0 – 57, with higher scores indicating better test performance. There is no categorical cut-off score.

#### User Experience Questionnaire

Acceptance of robot-assisted and iVR-based arm training was assessed using the German version of the User Experience Questionnaire (UEQ). For a detailed description, see^31^.

### Interventions

All patients received standard rehabilitation of one to two hours per day on five days a week, including physical therapy for improving mobility, neuropsychology and speech therapy if needed. No other upper limb rehabilitation than the one used in the study was provided to the study participants.

#### Robot-assisted arm training and VR-based arm training

Both intervention groups received a total of ten therapy sessions over a maximum period of three weeks. Group therapies were performed daily (with a maximum of four consecutive days without therapy) in groups of two to three patients under the guidance of an occupational therapist. Each therapy unit consisted of 20 minutes of conventional therapy and 20 minutes of robot-assisted or iVR-based arm training.

During conventional therapy, patients practiced movements of the affected arm such as reaching and grasping. Both isolated training on the hand, the elbow, and the shoulder as well as whole-arm training was applied. Although the training was mainly performed using the affected arm, some elements required both arms. Patients were assisted by a qualified therapist (hands-on) to mobilize elevated muscle tone, to initiate hand or arm movements, and/or to provide weight relief.

Robot-assisted training of the paretic arm was performed using the ARMEOSpring® system (Hocoma). Of note, some authors refer to the ARMEO system as electromechanical device rather than a robotic device; however, “robot-assisted training” is a commonly used term in the literature which was adopted for the ease of reading. The ARMEOSpring® consists of an ergonomically designed exoskeleton, which is attached to the paretic limb to provide anti-gravity support of the paretic arm through weight reduction. All movements have to be initiated and performed by the patient. The weight reduction can be adjusted for each individual patient, which allows them to move their paretic limb throughout its active range of motion. The exercises are displayed on a computer screen in front of the patient. The therapist can choose from a library of different functional tasks, which the patient can complete by themselves. Direct visual feedback on movement performance is provided on screen.

IVR-based arm training of the affected arm was performed using the CUREO® system (provided by CUREosity, www.cureosity.de) with a head-mounted display (Oculus Quest), two controllers, and a tablet. The controllers were used to record the movements of the hands allowing interaction with the virtual environment. Patients practiced gross motor movements of the affected arm on a table or starting from their lab using playful tasks with visual and auditory feedback. The first task consisted of a caterpillar that needed to be guided towards a fruit on a pre-defined path (see figure 1 in Supplemental Material). This path could take the shape of a simple line (either horizontal from left to right or vice versa or from front to back or vice versa), a triangle, square or circle. The width and depth of the shape could be adapted to the patients’ maximum range of movement. Depending on the patients’ severity of impairment, the task could be completed solely on the table or with the arm raised against gravity. To facilitate arm movements on the table, a towel could be placed under the arm to reduce friction. In the second task, meteorites flew towards the patient and had to be caught by “touching” the meteorites with the virtual hand. At a lower level, all meteorites could be caught by moving the affected hand across the table, while the range of motion depended on the patients’ maximum range of movement. At a higher level, the meteorites also flew well above table height, so that patients had to lift their hand in order to catch them.

### Statistical Analysis

Statistical analysis was performed using IBM SPSS (version 20) and R (version 4.3), aiming to answer five main questions (Q).

[Q1] First, changes in total ARAT scores (difference in scores after and before intervention) were tested for each group separately to test for intervention-related improvements.

[Q2] Second, robot-assisted and iVR-based arm training were compared based on the changes in total ARAT scores (ΔARAT; difference in scores after and before intervention) to test whether iVR-based training is not inferior compared to robot-assisted training (H_0_: iVR > robot-assisted -margin of non-inferiority; H_1_: iVR ≤ robot-assisted – margin of non-inferiority). To test for non-inferiority, a margin of non-inferiority of 3 points was considered, as this deviation represents the minimum changes in ARAT scores considered to be clinically relevant ^33^. [Q2.2] In addition, a post-hoc comparison of both intervention groups was planned in case a non-inferiority of iVR-based arm training was observed.

[Q3] Third, robot-assisted and iVR-based arm training were compared based on their clinical relevance. Clinical relevance was assessed depending on the prior ARAT scores ^33^, where an improvement of 3 points was required for baseline scores between 0 and 7, 4 points for initial scores between 8 and 13, 5 points for initial scores between 14 and 19, and 6 points for initial scores between 20 and 39.

[Q4] Fourth, the number of drop-outs was compared across interventions in case of occurrence.

[Q5] Fifth, user experiences were compared between interventions based on the UEQ scores.

Additional control analyses were performed to verify the randomization procedure by comparing intervention group with respect to age, gender, duration after stroke, affected body side, and ARAT scores prior to the intervention.

To test for statistical differences and improvements, the normality of the data was tested using the Shapiro-Wilk test. Non-inferiority was assessed by a 95% confidence interval, representing the range of the true difference between robot-assisted and iVR-based arm training. Here, inferiority is reflected by a confidence interval that do not exceed the margin of non-inferiority. The confidence interval was calculated using bootstrapping (1000 samples), as this procedure is not limited to a specific distribution of the data. Group-wise improvements in ARAT scores were tested using either one-sample t-tests or Wilcoxon signed-rank tests dependent on data normality. Group comparisons were performed using either parametric student’s t-tests or non-parametric Mann-Whitney-U-test. Nominal data, such as gender, affected body side, clinical relevance, and the number of drop-outs were compared between interventions using the Chi-squared test. Bonferroni correction for multiple comparisons was performed by multiplying the calculated p-values by the number of tests performed to answer each question, maintaining an interpretable alpha error of 5%. As this correction can yield p-values above 1, high p-values are indicated as “>1”. Data were reported as mean scores with corresponding standard errors or as median scores (M) with interquartile ranges (IQR) for normal or non-normal distributed data, respectively.

## Results

As the normality of all data was not fulfilled, only non-parametric statistics were applied. The median duration after stroke was 39 days for the robot-assisted group (IQR = 31.5 days) and 42 days in the iVR group (IQR = 33.5 days), respectively. In both groups, more patients suffered from stroke-related impairments of the left body side (robot-assisted group: 77%; iVR group: 53%).

Comparing participants across robot-assisted and iVR-based arm training, there were no differences in age (p_Mann-Whitney-U_ > 1), gender (p_Chi-squared_ > 1), duration after stroke (p_Mann-Whitney-U_ > 1) and affected body side (p_Chi-squared_ = .44) across groups. Furthermore, no difference in ARAT scores at baseline was observed (p_Mann-Whitney-U_ > 1) when comparing robot-assisted (M = 4; IQR = 13.5) and iVR-based arm training (M = 4; IQR = 6.5 (see also Table 1).

[Q1] Improvements in ARAT scores were found for both, the robot-assisted (p_Wilcoxon signed-rank_ = .002; ΔARAT score: M = 3; IQR = 7.5) and iVR-based arm training (p_Wilcoxon signed-rank_ < .001; ΔARAT score: M = 14; IQR = 12; see Figure 3).

[Q2] Based on the changes in total ARAT scores, no inferiority of robot-assisted arm training compared to iVR-based arm training was found, as the calculated confidence interval (4.5 to 17) do not fall below the determined margin of inferiority (see Figure 2). [Q1.2] Additionally, group comparison (p_Mann-Whitney-U_ = .004) indicates higher improvements after iVR-based (M = 14; IQR = 12) as compared to robot-assisted arm training (M = 3; IQR = 7.5; see Figure 3).

**Figure 2:**
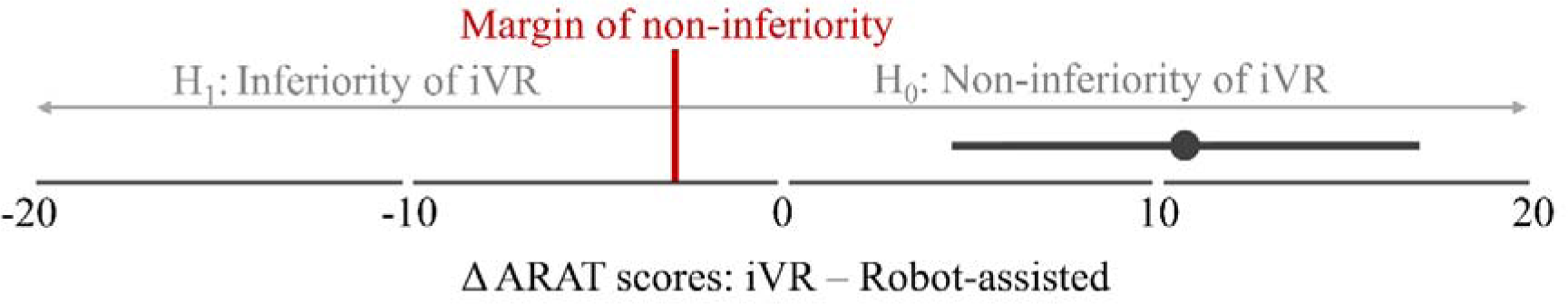
Difference in intervention-related changes in total ARAT scores. Confidence interval (95%) of the difference in intervention-related changes in total ARAT scores (difference in scores after and before intervention) between robot-assisted and iVR-based arm training. The margin of non-inferiority (3 point) is shown as red bar, indicating the decision criterion of the hypotheses tested (H0: iVR > robot-assisted - margin of non-inferiority; H1: iVR ≤ robot-assisted – margin of non-inferiority).

**Figure 3:**
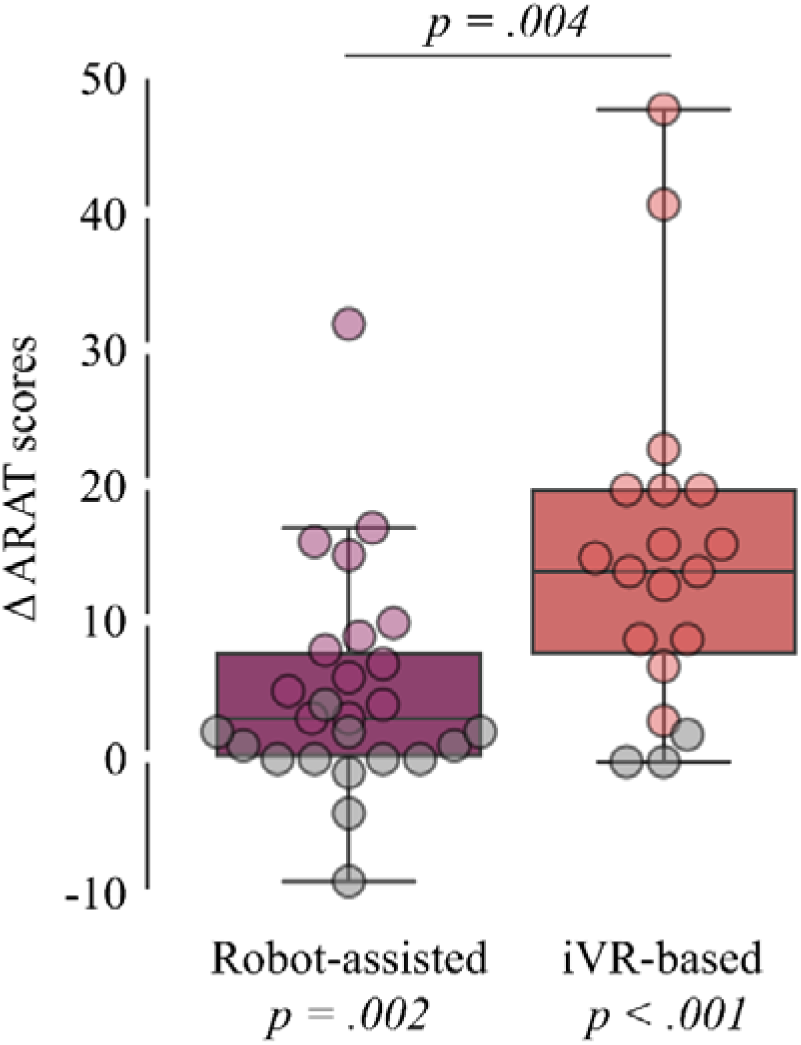
Intervention-related changes in total ARAT scores. Changes in total ARAT scores (difference in scores after and before intervention) for each intervention. Statistical p-values comparing changes in total ARAT scores against zero (bottom) and between interventions (top) are shown. Patients without clinically relevant improvement are shown in grey.

[Q3] In addition, a significant difference between the two interventions was observed with respect to the clinical relevance score: 84% in the iVR-group (16 out of 19) and 50% in the robot-assisted group (13 out of 26) showed clinically relevant improvements in the ARAT scores (p_Chi-squared_ = .036).

[Q4] Furthermore, no statistically significant difference in intervention-related drop-out rate was found (p_Chi-squared_ =.149) comparing iVR-based (5 out of 24) and robot-assisted arm training (2 out of 28).

[Q5] Comparing user experiences between the two types of interventions (UEQ; see Figure 4), no differences were found regarding attractiveness (p_Mann-Whitney-U_ = .45), perspicuity (p_Mann-Whitney-U_ > 1), efficiency (p_Mann-Whitney-U_ > 1), dependability (p_Mann-Whitney-U_ > 1), stimulation (p_Mann-Whitney-U_ = .582), and novelty (p_Mann-Whitney-U_ > 1). For more details, see Table 2.

**Figure 4:**
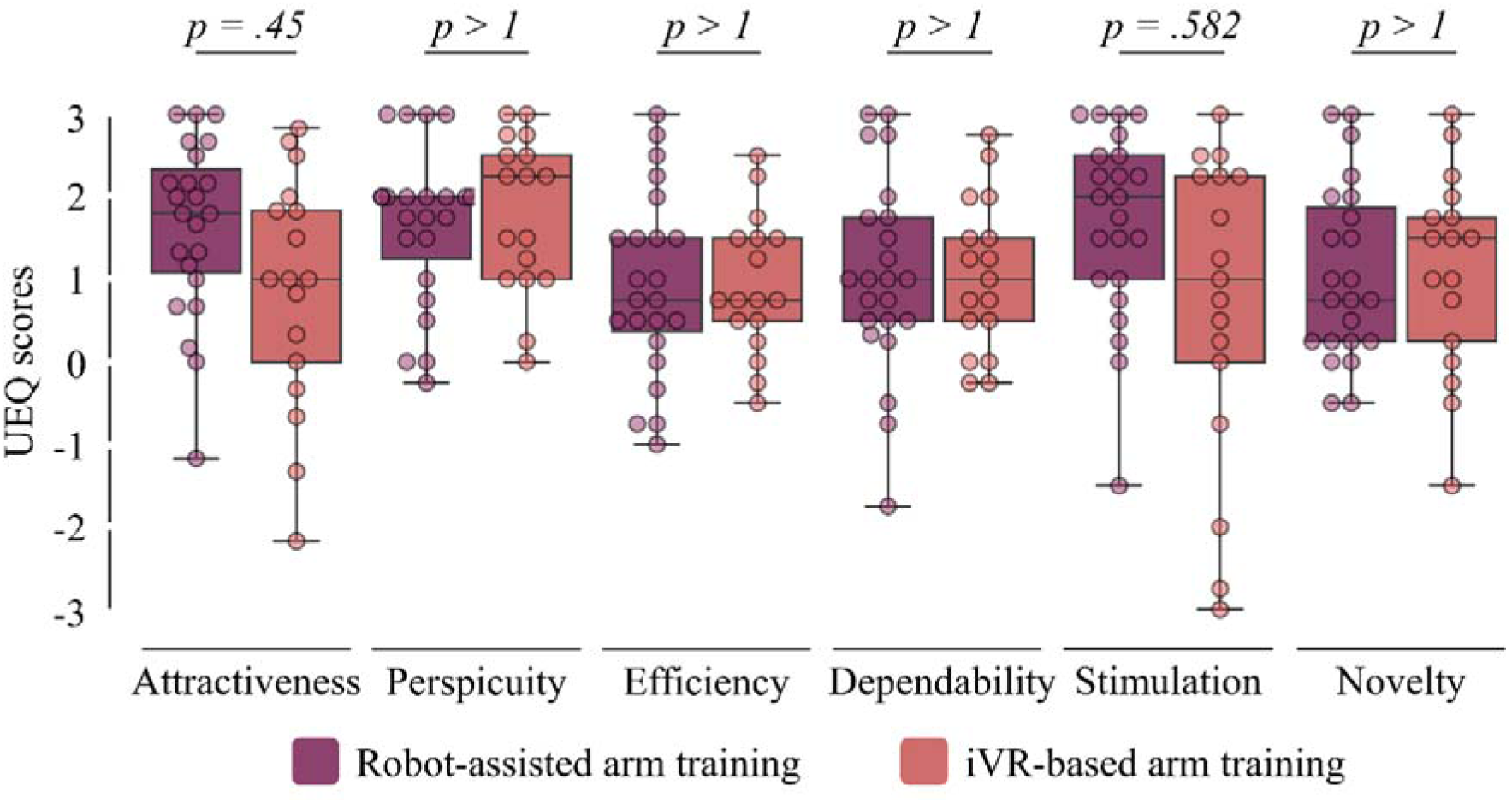
User experiences. Scores of the user experiences of all six items from the User Experience Questionnaire (UEQ) for robot-assisted (violet) and iVR-based arm training (red). Statistical p-values comparing experience ratings between interventions are shown (top).

**Table 2:** User Experience Questionnaire. User experience ratings for robot-assisted and iVR-based arm training as median scores (M) with corresponding interquartile ranges (IQR). Using the User Experience Questionnaire (UEQ), both interventions were rated on six scales (attractiveness, perspicuity, efficiency, dependability, stimulation, novelty), each ranging from −3 (not applicable) to +3 (fully applicable).

|  | Attractiveness |  | Perspicuity |  | Efficiency |  | Dependability |  | Stimulation |  | Novelty |  |
| --- | --- | --- | --- | --- | --- | --- | --- | --- | --- | --- | --- | --- |
|  | IQR |  | IQR |  | IQR |  | IQR |  | IQR |  | IQR |  |
| Robot-assisted training | .8 | .25 | .0 | .75 | .8 | .13 | .0 | .25 | .0 | .50 | .8 | .63 |
| VR-based training | .0 | .83 | .3 | .50 | .8 | .00 | .0 | .00 | .0 | .25 | .5 | .50 |

## Discussion

Both robot-assisted arm trainings using the ARMEOSpring® ^34–36^ as well as VR-based arm trainings (non-, semi- and immersive) ^21–23^ have been shown to be beneficial add-ons to conventional therapy in the treatment of an arm paresis after stroke. In the present study, we compared a newly developed iVR-based upper limb training (CUREO®) with a robot-assisted arm training (ARMEOSpring®) in a population of subacute stroke patients with severe arm paresis in a randomized-controlled trial. Both technology-based treatments were delivered as time-matched trainings as part of the standard upper extremity training which consisted of twenty minutes of conventional task-oriented training and twenty minutes of either robot-assisted or iVR-based training.

The study was based on the hypothesis that the newly developed iVR-based training would be at least as effective in improving upper limb activity in stroke patients as compared to the established ARMEOSpring® training. At a group level, we found that both interventions were associated with significant improvements of motor activity of the affected arm as measured with the ARAT with more extensive improvements in the iVR-group as compared to the robot-assisted group. No inferiority of iVR-based training compared to the robot-assisted arm training could be detected, post hoc group comparison even indicated higher improvements in the iVR group. When evaluating clinical relevance of individual improvements as suggested by Rodgers and colleagues (2019), we found that 84% of patients treated with the CUREO® and 50% of the patients treated with the ARMEO® showed a clinically relevant improvement of arm function. This difference was statistically significant and could neither be attributed to baseline performance nor to differences in the total amount of therapy provided. Furthermore, the additional beneficial effect of the iVR-based training was unlikely to be caused solely by motivational effects (gamification in a three-dimensional environment) as we did not find significant differences in user acceptance.

Both systems used in the current study apply high end technologies to improve arm function and activity. The systems differ with regards to two major aspects: immersion and anti-gravity support. The CUREO® system stages an immersive type of VR, which fully surrounds the user providing multisensory stimulation in a three-dimensional environment. The high degree of immersion, which is defined as the sensory fidelity a VR-system reaches ^20^, allows the user to feel present in and to be part of the virtual environment. In contrast, the ARMEO® uses non-immersive VR on a computer display. In both systems, users interact with the virtual environment, allowing a high sense of agency, i.e. the subjective feeling of having control over the avatar’s movements and actions. A recent meta-analysis showed that iVR-based trainings were superior to non-immersive and semi-immersive programs in the treatment of arm paresis after stroke ^23^. This beneficial effect of iVR-based training has been associated with perceived body ownership over the avatar’s arm in the virtual environment as well as a high sense of agency. The more the patient perceives the virtual avatar arm as part of their own body, the better the therapeutic outcome of iVR-based arm trainings ^37^. Moreover, visual characteristics of the virtually displayed arm significantly influence the users’ perception of their physical abilities and, consequently, their behaviour. For instance, it has been shown that perceiving the virtual arm as stiff and cold negatively affects the users’ speed of movement and sense of agency ^38^. Thus, it is possible that body ownership over a healthy virtual arm as displayed in the VR environment improved patients’ perception of their physical abilities and helped them to perform / initiate movements, which they would have thought impossible with their real, highly paretic arm. Moreover, it is conceivable that patients in the iVR-group experienced a high degree of sense of agency over their virtual arm, which, in turn, might have led to a higher extent of integration of the arm into the newly evolving body image and, on a neurobiological level, to a more effective integration in the neural circuitry that promotes functional recovery of the paretic arm. Future studies need to include a measure for both body ownership and sense of agency and investigate more closely how the perception of the virtual arm affects the patients’ physical abilities.

Secondly, in contrast to the CUREO®, the ARMEOSpring® provides anti-gravity support of the paretic arm. The ARMEOSpring® assists self-initiated movements during training through weight support, which allows patients with severe arm paresis to perform movements against gravity even if they are not (yet) capable of lifting or holding the paretic arm against gravity during arm movements in three-dimensional space. This allows a higher number of movement repetitions and an increase of training intensity, which has been shown to reduce upper limb impairment after stroke ^15,33^. The CUREO® system, on the other hand, does not provide anti-gravity support. At the lowest level all movements are performed in the horizontal plane with the arm placed on a table, while the range of motion required for successful completion of the task is adapted to the patients’ physical abilities. Patients who are able to overcome gravity, at least to some extent, are challenged to perform a more difficult task, where movements are not confined to the horizontal plane. This might drive the development of adaptive strategies to complete the task without following the ideal and/or previously (before stroke) used arm trajectory.

In the present study, we evaluated the efficacy of both a robot-assisted and an iVR-based training using the ARAT, which assesses upper extremity function at the level of activity and limitations rather than at the level of structure/function and impairments (following ICF nomenclature). The ARAT includes several tasks, where patients have to lift the paretic arm against gravity (see methods section). Thus, one might have expected that patients who trained movements in the three-dimensional space with weight support using the ARMEO® would show a higher extent of improvement. However, in this study, patients who trained with the CUREO® reached higher ARAT scores, possibly due to the fact that they had to adapt strategies to fulfil the tasks without relying on additional support. Earlier studies have repeatedly demonstrated that robot-assisted arm trainings reduce upper limb impairment after stroke. However, this might not necessarily translate into the improvement of upper limb activity ^15,33^. Against this background, it has been proposed that robot-assisted arm trainings should be used to reduce upper limb impairment through intensive and highly repetitive arm trainings, especially at the beginning of treatment in severely affected patients, accompanied/followed by conventional therapy, which then targets upper limb limitations in the activities of daily living, once a certain level of recovery has been achieved ^19^.

### Limitations

The study sample is rather small and larger studies will be needed to confirm the observed effects. More importantly, the study lacks a control group, that received dose-matched conventional therapy. Therefore, we cannot make any conclusions about the effects of iVR-based training in comparison to dose-matched conventional therapy. Earlier randomized-controlled studies on the efficacy of iVR-based arm trainings showed that the delivery of practice, i.e. whether the exercises were displayed and performed in iVR (unsupervised) or in a standard occupational therapy setting (in a dose-matched manner), did not significantly affect the outcome, in that both groups improved equally ^26–28^. Thus, it has been argued that the benefit of including iVR-based trainings in neurorehabilitation lies primarily in the possibility to increase training intensity (time and/or frequency) by translating established therapeutic concepts into iVR to provide unsupervised self-training. In this context, it has been shown that VR is effective as a therapeutic tool, when it promotes basic principles of neurorehabilitation, namely task-specific practice, explicit and implicit feedback, increasing levels of difficulty, variability of practice and mechanisms to promote the use of the paretic limb ^39^.

Furthermore, our study is based on a single outcome measure, i.e. the ARAT. The ARAT is an instrument designed to measure arm activity, as such we did not measure recovery from arm impairment at the level of structure and function, which might have yielded different results. Neurorehabilitation primarily aims at the increase of arm activity, however, it is well conceivable that patients at different stages of recovery profit from different strategies, i.e. highly repetitive, anti-gravity supported training in the early phase after stroke with the aim to support movement initiation and to reduce impairment with a follow-up of activity-oriented training. More research with different assessment tools and different subgroups of patients is needed to develop fine-grained strategies to address the different stages and dynamics of recovery.

Finally, our study does not allow any conclusions on the neural mechanisms by with iVR- or robot-assisted trainings unfold their therapeutic effects; as such, our study is purely descriptive. As discussed above, body-ownership and sense of agency are interesting concepts that might mediate the beneficial effects of immersive and non-immersive VR on recovery. However, this needs to be investigated in more detail in future studies.

### Conclusion and outlook

The present study showed that the newly developed iVR-based arm training CUREO® was not inferior to the robot-assisted arm training using the ARMEOSpring® in improving upper limb activity in subacute stroke patients with severe arm paresis. 84% of patients who trained with the iVR-based system in combination with conventional therapy showed a clinically relevant improvement of arm activity after three weeks of treatment. As such, iVR-based devices have the potential to play a future role in the rehabilitation of stroke patients with severe arm paresis.

Moreover, iVR generally allows the continuation of training after hospital discharge when used in the conceptual framework of tele-rehabilitation. This is particularly relevant as stroke patients benefit from a continuation of therapy at home which can be accomplished by iVR-based training ^28^ embedded in a rather smooth transition from the rehab facility to patients’ home.

Future studies should include measures for both motor function and activity in order to examine whether different types of technology-supported trainings have differential effects on motor recovery. From a mechanistic perspective, it will be of great interest to better understand the neural basis of immersion and how it might be modulated to yield better therapeutic outcomes. As such, it will be worth investigating whether the display of a healthy or even strong arm in an immersive virtual environment affects the users’ perception of their physical abilities, which, in turn, might have an additional beneficial effect on motor recovery.

## Declarations

### Ethics Approval and Consent to Participate

The study was conducted in accordance with the Declaration of Helsinki and had been approved by the Ethics Committee of the Ärztekammer Nordrhein (serial number: 2020079). Prior to study enrolment, participants gave written informed consent.

### Consent for Publication

Prior to study enrolment, participants gave written informed consent for publication.

### Author contributions

**Kira Lülsdorff**: Conceptualization, Writing – original draft

**Frederick Junker**: Formal analysis, Visualization

**Bettina Studer**: Conceptualization, Methodology, Supervision, Writing – review & editing

**Heike Wittenberg**: Conceptualization, Methodology, Writing – review & editing

**Heidrun Pickenbrock**: Conceptualization, Methodology, Project administration, Resources, Supervision, Writing – review & editing

**Tobias Schmidt-Wilcke**: Conceptualization, Methodology, Project administration, Resources, Supervision, Writing – review & editing

## Data Availability

All data produced in the present study are available upon reasonable request to the authors.

## Acknowledgements

We would like to acknowledge CUREosity GmbH for providing two iVR systems for the study.

## Availability of data and material

Not applicable.

## Abbreviations

ARAT: Action Research Arm Test
BBT: Box and Block Test
HMD: head mounted display
ICF: International Classification of Functioning
iVR: immersive virtual reality
UEQ: User Experience Questionnaire
VR: virtual reality

