## Supplemental Material for "Neurorehabilitation of the upper extremity – Immersive virtual reality vs. robot-assisted training. A comparative study"

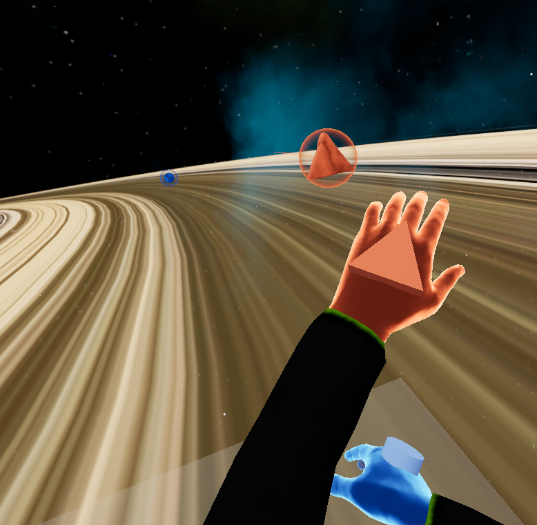

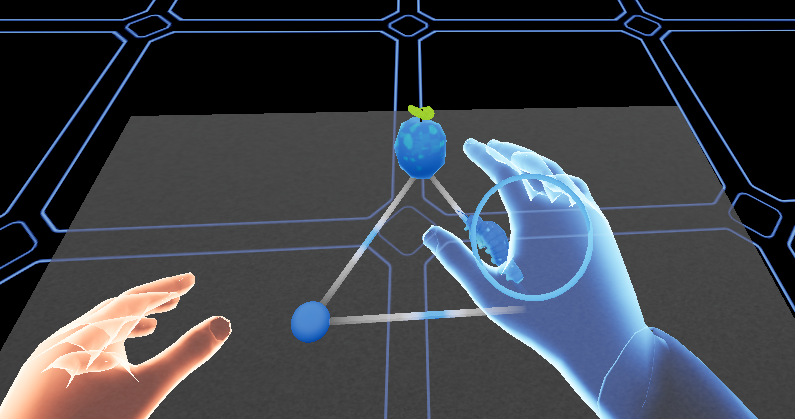

**Figure 1:** The iVR-based unilateral arm training. On the left, a caterpillar has to be guided along a pre-defined path towards a fruit. On the right, moving meteorites have to be caught by “touching” them with the affected hand.
